# Carbapenem Resistance Among Gram-Negative Bacteria in Bioaerosols at Biomedical Waste Disposal Site in Ahmedabad, India

**DOI:** 10.64898/2026.10.22.26366283

**Authors:** Ram Shankar Barai, Sanjana Ravat, Pankaj Barfal, Mugdha Tiwari, Gitika Kharkwal, Nitika Garg, Paltu Kumar Dhal, Snehal Chavan, Gargi Das, Santasabuj Das

## Abstract

Antimicrobial resistance (AMR) is a global public health concern, with Gram-negative carbapenem-resistant bacteria (CRG) classified by WHO as critical priority pathogens. Biomedical waste disposal sites (BMWs) may act as reservoirs for resistant organisms, posing occupational and community risks. This study investigated the phenotypic and genotypic characterization of Gram-negative carbapenem resistant bacteria in bioaerosols collected from BMW facilities in Ahmedabad, Gujarat, India.

Bioaerosols were sampled using active Six-stage Andersen Impactors and passive settled plates. Isolates were identified by MALDI-TOF, followed by antimicrobial susceptibility testing (AST). Whole genome sequencing (WGS) was performed for 26 representative isolates, and resistance gene profiling was conducted using CARD and ResFinder databases.

Representative isolates belonged to Moraxellaceae, Enterobacteriaceae, and Pseudomonadaceae families. All species showed resistance to linezolid, and approximately 60% were multidrug resistant to carbapenems, cephalosporins, and fluoroquinolones. Genotypic analysis confirmed the prevalence of carbapenemase genes, with blaNDM-1 most frequent, followed by blaNDM-7, blaVIM-2, blaVIM-11, blaOXA-23, and blaOXA-235. This is the first report documenting Acinetobacter baumannii, A. schindleri, A. towneri, Klebsiella pneumoniae, and Pseudomonas juntendi in bioaerosols from BMW sites.

Biomedical waste environments serve as critical reservoirs of AMR. These findings emphasize the need for surveillance, protective measures, and integration of environmental AMR monitoring into public health policy.

## Introduction

Antimicrobial resistance (AMR) is the major public health concern of the 21^st^ century ^1^. By 2050, AMR is projected to contribute to approximately 5 million deaths annually, with the majority occurring in low and middle income countries ^1,2^. Carbapenems, a class of beta-lactam antibiotics exhibits broad-spectrum activity against a wide range of Gram-positive and Gram-negative bacterial pathogens, making them effective in the treatment of bacterial infections ^3^. Carbapenem resistance Gram-negative bacteria (CRG) included in the critical group of WHO bacterial priority pathogens ^2^.

Healthcare activities generate biomedical waste of which 10-15% is infectious material ^4^. Despite appropriate packaging of infectious biomedical waste at healthcare facilities prior to disposal, the growing volume of biomedical waste (BMW), combined with suboptimal handling, transportation, storage and management practices, may contribute to the environmental spread of AMR ^5^. Air is the largest microbial ecosystem and bioaerosols act as a transport medium for microbial content at Biomedical Waste Disposal Site (BMW) ^6^. CRGs in bioaerosols may directly spread infection or serve as a reservoir of resistance genes, thereby elevating the risk of resistance transmission to other pathogenic microorganisms and posing a significant threat to human health ^7^. The complex microbial composition in bioaerosols may facilitates gene transfer, including the exchange of antibiotic resistance genes (ARGs) in bacterial species within BMW storage and disposal sites ^8,9^.

Hospital based study in India provided the prevalence of carbapenem resistance genes and coexistence of multi drug resistance in Gram-negative bacteria ^10^. Most of the AMR studies in India focus on hospital and data is scarce on ARBs in bioaerosols at BMW in India.

Environment is an important component that has wide reaching effects at community level, and thus the present study aimed to evaluate the AMR burden in the environment of the BMW ^11^. This cross-sectional study was conducted to identify airborne CRG isolates from BMW in Ahmedabad, Gujarat, India and to characterize their phenotypic and genotypic resistance patterns.

## Materials and Methods

### Air Sampling and Bacterial Isolation

The BMW have several functional subunits or plants like incinerator, autoclave, temporary storage area, segregation and shredding plant and administrative office. Bioaerosol samples were collected from all the functional subunits of BMW in September 2022-2024 under the Network Program on Antimicrobial Resistance, Superbugs, and One Health at Ahmedabad, Gujarat, India by the following methods ^12^.

#### 1. Passive sampling (Settled plate method)

Passive air sampling was done by exposing nutrient agar and MacConkey agar plates at ground level for 10 minutes at each functional subunit. Isolates obtained from these plates were assigned location-specific codes to identify their source within the BMW facility.

The codes were: F – incineration unit, G – autoclave unit, H – storage area, I – shredder unit, J – segregation area, and K – control site located near the administrative office.

#### 2. Active sampling (Six-stage Andersen Impactors)

Active air sampling was carried out using a six-stage Andersen cascade impactor (Andersen Instruments Inc., USA), which was operated at a calibrated flow rate of 28.3 L/min. The impactor was positioned at a height of 1.2 m above the ground level to simulate the human breathing zone and was exposed for a duration of 3 minutes at each selected unit within the BMW.

Each stage of the cascade impactor was equipped with an agar-containing plate and featured progressively smaller jet nozzles designed to create increasing impaction velocities. This configuration enabled aerodynamic size-based segregation of airborne particles. Aerosols entered from the top and passed sequentially through the stages, depositing particles by size: stage 1 collected particles >7□µm, stage 2 collected 7–4.7□µm, stage 3 collected 4.7–3.3□µm, stage 4 collected 3.3–2.1□µm, stage 5 collected 2.1 –1.1□µm, and stage 6 collected 1.1–0.65□µm particles ^13^. Sampling locations were coded to identify the origin of each isolate within the BMW facility: A – incineration unit, B – autoclave unit, C – storage area, D – shredder unit, and E – control site located outside the BMW premises.

After sampling, inoculated plates were incubated at 37 °C for 12–18 hours to facilitate the development of colony-forming units (CFUs). Distinct colonies were isolated and purified through repeated sub-culturing using the four-quadrant streaking technique. Pure bacterial cultures were preserved as 20% glycerol stocks and stored at –80□°C for long-term preservation.

### Bacterial identification via MALDI-TOF

A single colony from the incubated pure culture agar plate was aseptically transferred onto an MPT-384 ground steel target plate for MALDI-TOF MS identification. (Bruker Daltonics, Bremen, Germany). Colony lysis was performed by adding 1□µL of 70% formic acid (Merck Life Sciences, India). After air drying, 1□µL of matrix solution; composed of 50% acetonitrile, 2.5% trifluoroacetic acid (both from Merck Life Sciences, India), and 10 mg/mL α-cyano-4-hydroxycinnamic acid (Sigma-Aldrich, St. Louis, USA) was added to each lysed sample. Following a second air drying step, the samples were subjected to MALDI-TOF MS analysis using the Autoflex maX system (Bruker Daltonics, Bremen, Germany). Mass spectra were acquired using FlexControl software and analyzed with MBT Compass (Bruker Daltonics, Bremen, Germany) for bacterial species identification ^14^.

### Antimicrobial Susceptibility Testing (AST) and Minimum Inhibitory Concentration (MIC) of Isolates

All Gram-negative bacterial isolates were subjected to AST using the Kirby–Bauer disk diffusion method on Mueller–Hinton agar (HiMedia, India), following the standard protocol recommended by the Indian Council of Medical Research (ICMR, 2022) for antimicrobial resistance surveillance^15^ (Table 1).

**Table 1:**
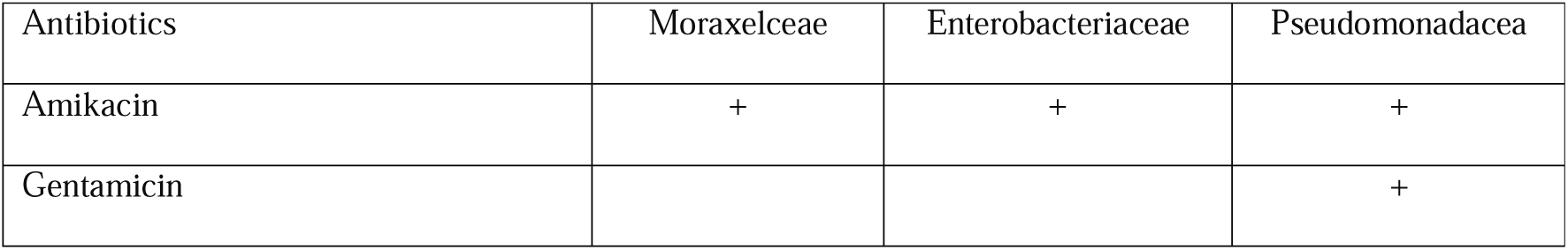
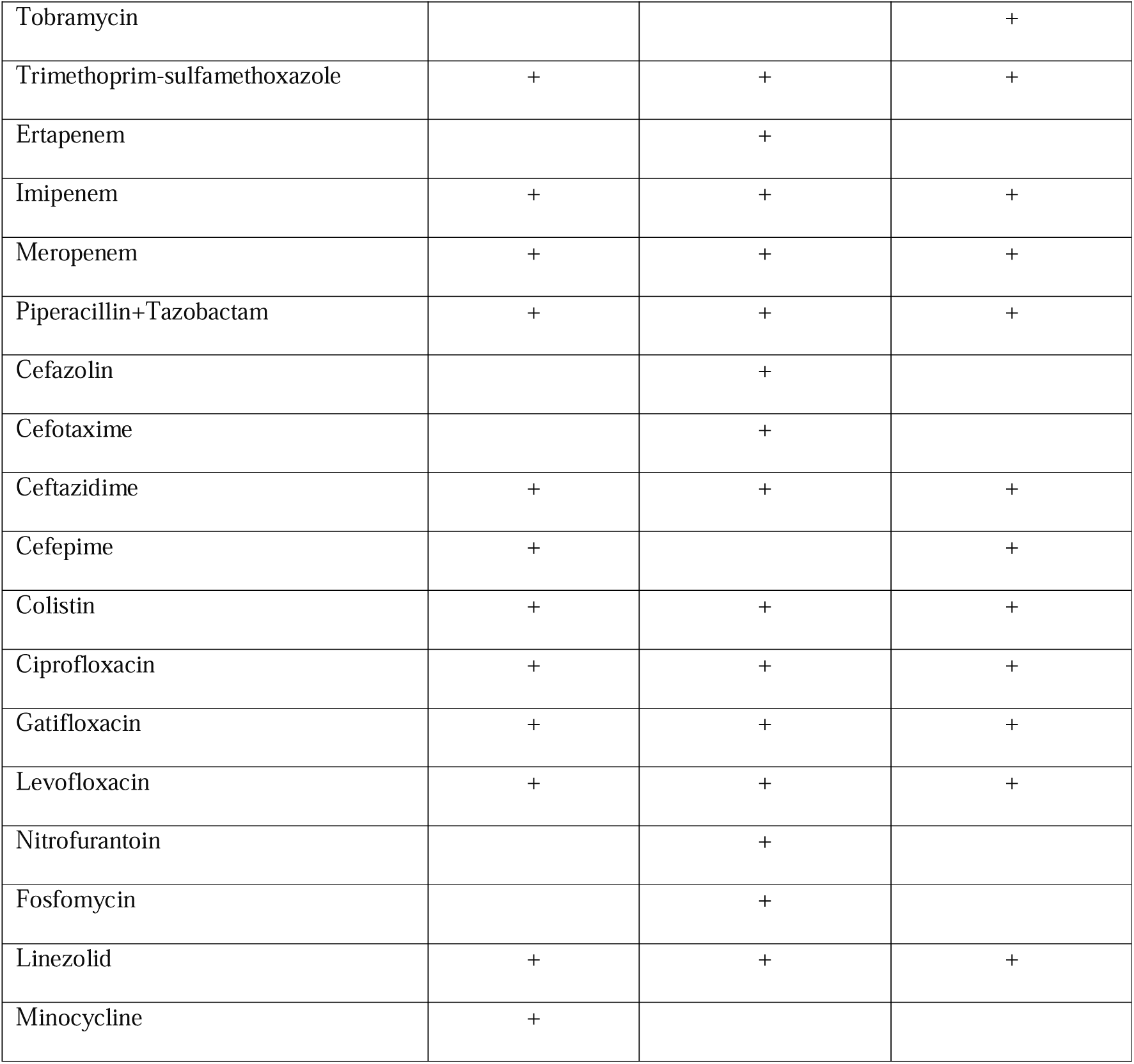
List of antibiotics used for AST and MIC profiling as per ICMR guidelines.

| Antibiotics | Moraxelceae | Enterobacteriaceae | Pseudomonadacea |
| --- | --- | --- | --- |
| Amikacin | + | + | + |
| Gentamicin |  |  | + |
| Tobramycin |  |  | + |
| Trimethoprim-sulfamethoxazole | + | + | + |
| Ertapenem |  | + |  |
| Imipenem | + | + | + |
| Meropenem | + | + | + |
| Piperacillin+Tazobactam | + | + | + |
| Cefazolin |  | + |  |
| Cefotaxime |  | + |  |
| Ceftazidime | + | + | + |
| Cefepime | + |  | + |
| Colistin | + | + | + |
| Ciprofloxacin | + | + | + |
| Gatifloxacin | + | + | + |
| Levofloxacin | + | + | + |
| Nitrofurantoin |  | + |  |
| Fosfomycin |  | + |  |
| Linezolid | + | + | + |
| Minocycline | + |  |  |

AST specifically targeted carbapenem antibiotics, including ertapenem (10 µg) imipenem (10 µg), meropenem (10 µg). Bacterial suspensions were prepared from overnight cultures and adjusted to 0.5 McFarland turbidity. The inoculum was evenly spread across MHA plates using a sterile swab, and carbapenem discs were placed aseptically on the agar surface. Plates were incubated at 37 °C for 16–18 hours, after which the zones of inhibition were measured and interpreted according to ICMR and CLSI interpretive standards.

Isolates exhibiting resistance or intermediate susceptibility to any of the tested carbapenems were further analyzed for minimum inhibitory concentration (MIC) values. MIC testing was performed using broth microdilution or E-test strips, depending on availability, across a range of concentrations as applicable to the resistance phenotype.

The resulting zone diameters and MIC values were entered into WHONET software for comprehensive antimicrobial resistance (AMR) profiling and trend analysis ^16^.

### Whole Genome Sequencing

Genomic DNA was extracted from 26 sub-cultured gram-negative ARB isolates using the CTAB method ^17^. DNA libraries were prepared using the QIAseq FX DNA Library Preparation Kit (Qiagen), according to the manufacturer’s protocol. Whole-genome sequencing was performed using Illumina NovaSeq 6000 platform. Raw sequencing reads were assessed for quality using FastQC version 0.12.1 ^18^. The low quality reads and adapter were removed by fastp version 0.23.4 ^19^. De novo assembly of paired-end sequencing reads was performed using SPAdes v3.15.5 genome assembler ^20^, and subsequently genome annotation was carried out with the NCBI Prokaryotic Genome Annotation Pipeline (PGAP. PGAP uses 2 pass of GeneMarkS+ tool ^21^ and hidden semi-Markov model (HSMM) of prokaryotic genome as template for homology based gene prediction available on github (https://github.com/ncbi/pgap.git) ^22^. The assembled contig sequences were submitted to the NCBI BioProject database Accession No. PRJNA1085497 for public access.

### ARG and Resistance associated mutation identification

To identify antimicrobial resistance genes (ARGs) from genome assemblies, three widely recognized bioinformatics tools were employed: The Comprehensive Antibiotic Resistance Database (CARD) Resistance Gene Identifier (RGI), ResFinder, and NCBI AMRFinderPlus. CARD RGI was used to predict resistomes by comparing input DNA (genome assemblies, metagenomes) or protein sequences against its antimicrobial resistance ontology using homology-based approaches ^23^. ResFinder was utilized to detect resistance determinants based on sequence similarity to a curated database of reference ARGs, facilitating the prediction of resistance phenotypes ^24^. NCBI AMRFinderPlus, a tool integrating a curated database of ARGs and resistance-associated mutations, was used to identify both acquired resistance genes and chromosomal mutations linked to antimicrobial resistance coupled with NCBI BLAST+ and custom PERL script ^25^.

The application of multiple tools allows cross-validation and increased accuracy in ARG detection. Identified ARGs and mutations were then correlated with antimicrobial susceptibility testing (AST) data to evaluate the concordance between genotypic and phenotypic resistance profiles, providing insights into the resistance mechanisms present in the studied isolates. The complete methodology is represented in the Figure-1.

**Fig.1:**
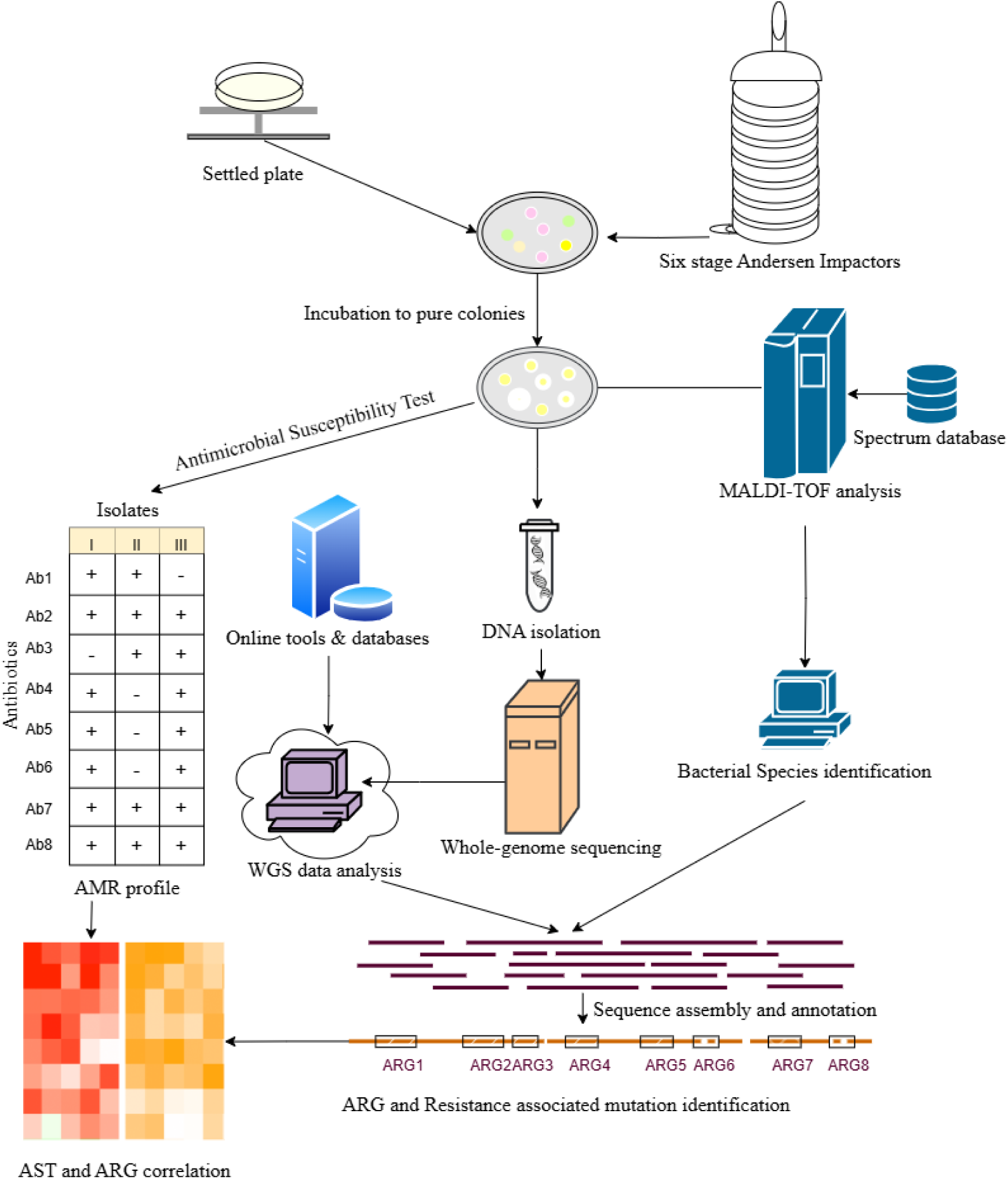
Workflow of the study.

## Results

### Profile of Gram-negative Bacterial Isolates

A total of 26 Gram-negative antimicrobial-resistant (AMR) isolates were characterized in this study, encompassing 7 strains from Moraxellaceae, 11 from Enterobacteriaceae, and 8 from Pseudomonadaceae. Species identification was performed using both MALDI-TOF and WGS. The two approaches demonstrated high concordance, with approximately 90% agreement at the genus level and ∼50% at the species level. The isolates were classified into 20 distinct species, primarily of environmental origin, but clinically relevant due to their multidrug-resistant phenotypes and potential to cause opportunistic infections. Detailed information along with the NCBI accession numbers corresponding to each WGS-verified isolate are provided (Table 2).

**Table 2:**
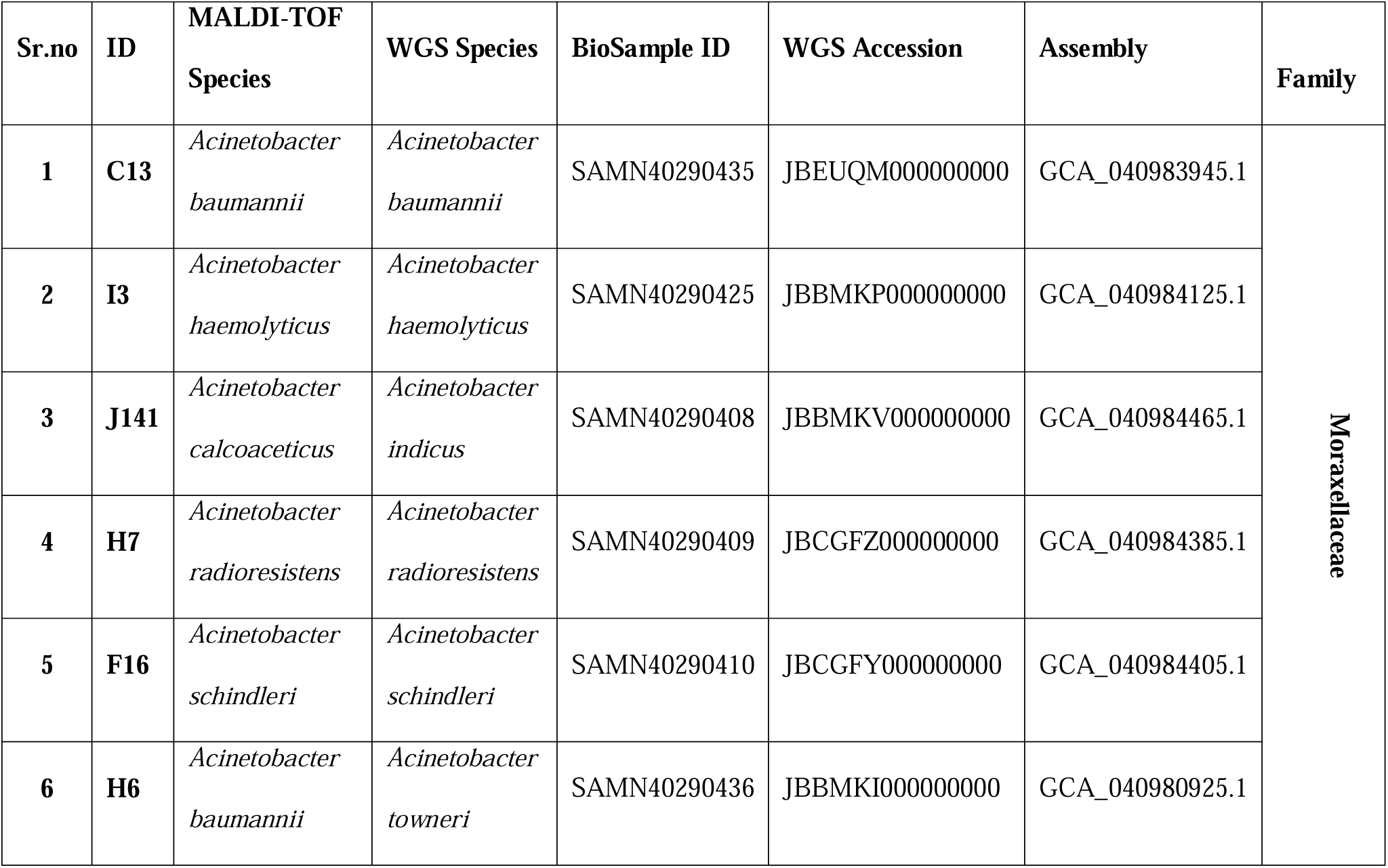
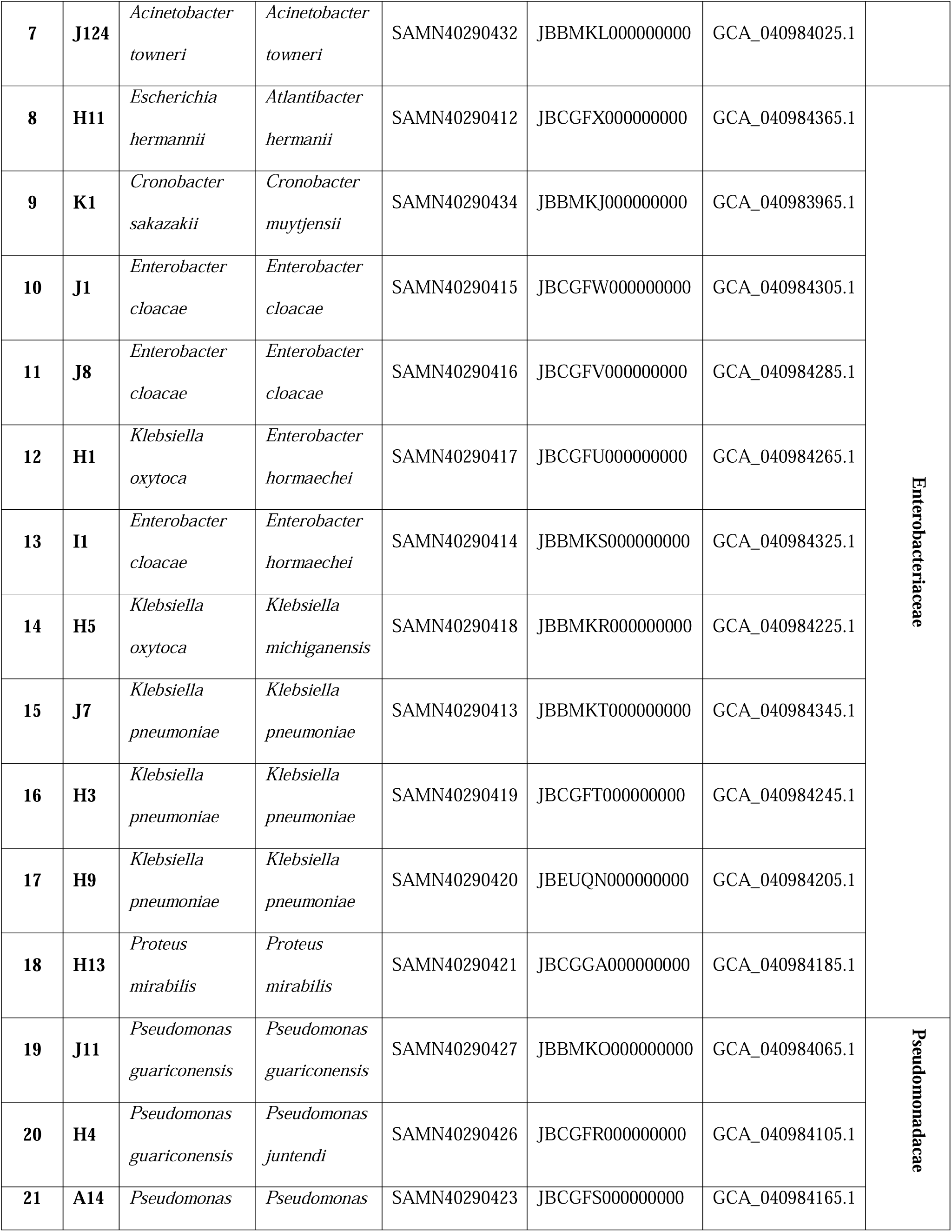
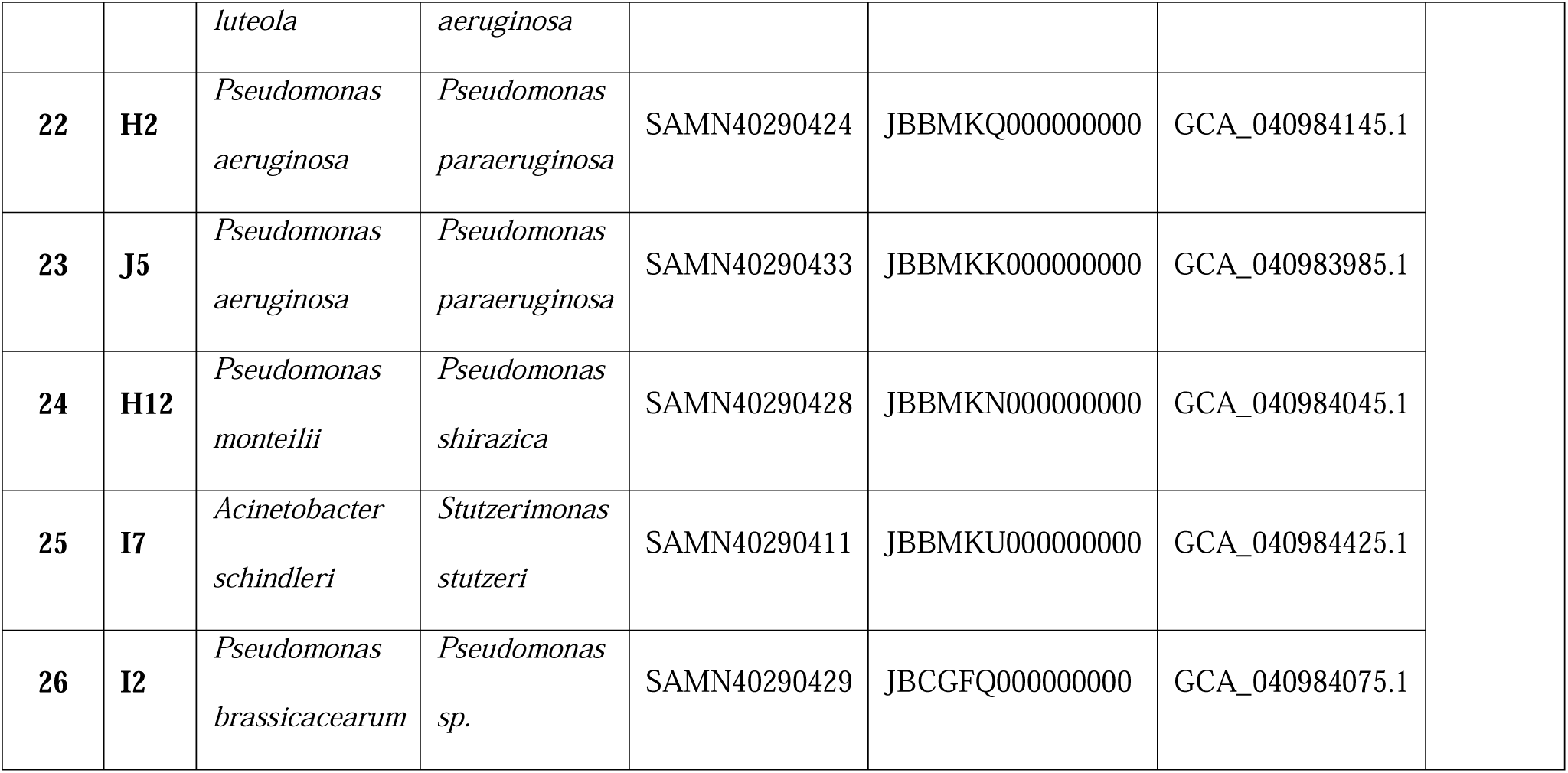
List of antimicrobial-resistant bacterial Isolates.

### Antibiotic Susceptibility Testing(AST) profile

AST of the Gram-negative isolates revealed a diverse range of resistance across 20 antibiotics (Figure 2). The highest rates of resistance were observed against cefazolin (100%), linezolid (96.15%), and cefepime (86.67%), indicating limited efficacy of these agents in the tested in tested isolates. Moderate levels of resistance were noted among fluoroquinolones, with ciprofloxacin (65.38%), levofloxacin (53.85%), and gatifloxacin (38.46%) displaying reduced antibiotic activity.

**Fig. 2:**
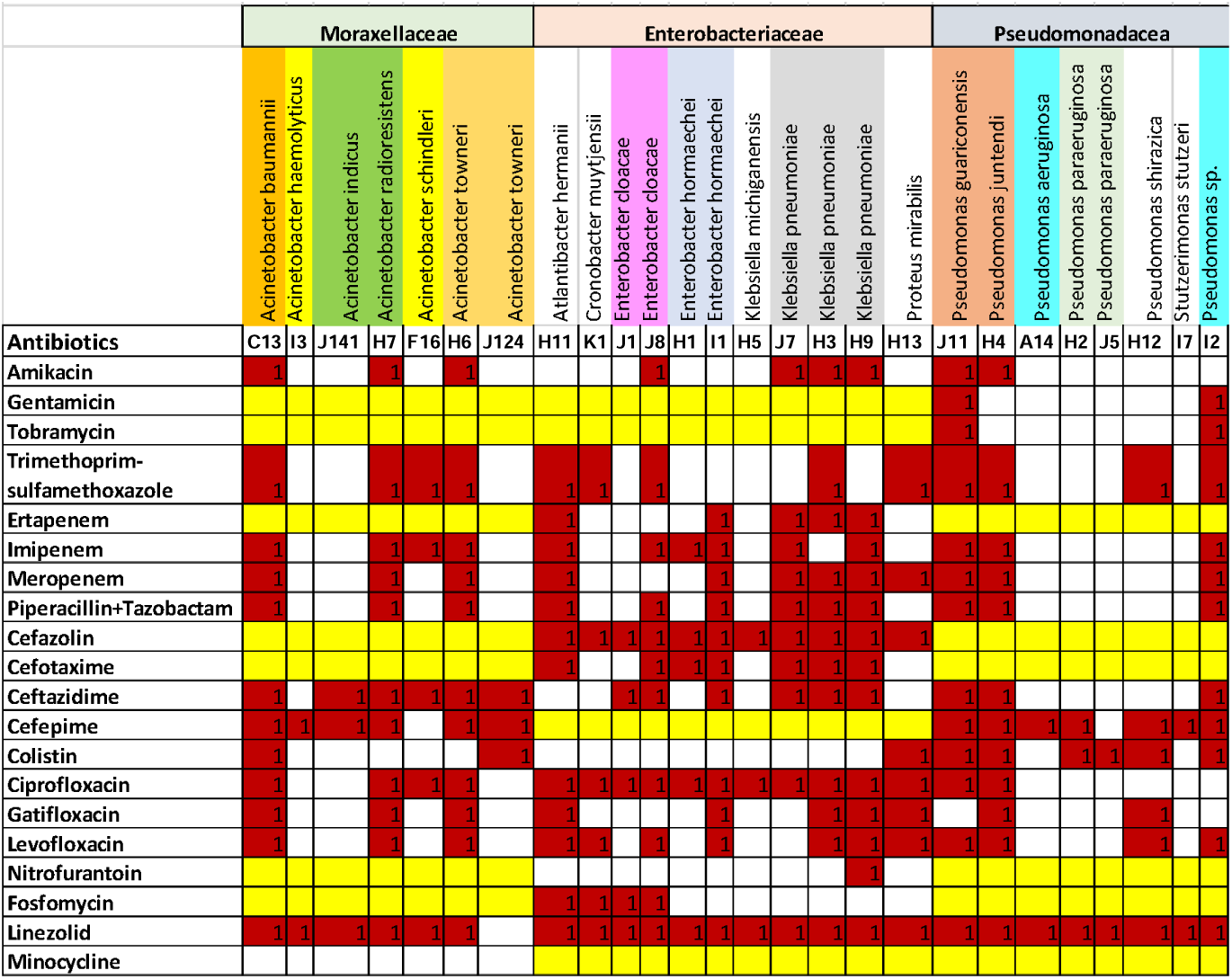
Heat map of 26 isolates showing Antimicrobial susceptibility test result.

Aminoglycoside antibiotics including amikacin (34.62%), gentamicin (25%), and tobramycin (25%) showed relatively higher effectiveness, while colistin, considered a last-resort agent, showed only 34.62% resistance, highlighting its waning utility.

Among the β-lactam class, cephalosporins showed variable activity; piperacillin–tazobactam (46.15%), cefotaxime (63.64%) and ceftazidime (57.69%) demonstrated moderate to high resistance, whereas cefepime exhibited the highest resistance among them. Carbapenem antibiotics ertapenem against Enterobacteriaceae while imipenem, and meropenem were tested against all the isolates to evaluate resistance patterns. The results revealed notable resistance across all three agents. 57.7% isolates were carbapenem-resistant out of them 42% expressed resistance for multiple carbapenems. Resistance for imipenem (50%), meropenem (46.15%), ertapenem (45.45%), and were modest, underscoring the growing prevalence of carbapenem-resistant strains.

Among the other agents tested fosfomycin (36.36%), trimethoprim–sulfamethoxazole (50%), and nitrofurantoin (9.09%) demonstrated comparatively higher susceptibility rates. Notably, minocycline exhibited complete susceptibility across all tested isolates.

### Genotypic Confirmation of Carbapenem resistance

Genotypic confirmation of carbapenem resistance was performed for 26 bacterial isolates using three genomic databases: RGI-CARD, ResFinder, and AMRFinderPlus (Table 3). A total of 29 resistance-associated genes were identified. Among the carbapenemase genes, blaNDM-1 was the most prevalent, followed by blaNDM-7, blaVIM-2, blaVIM-11, blaOXA-23, and blaOXA-235.

**Table 3:**
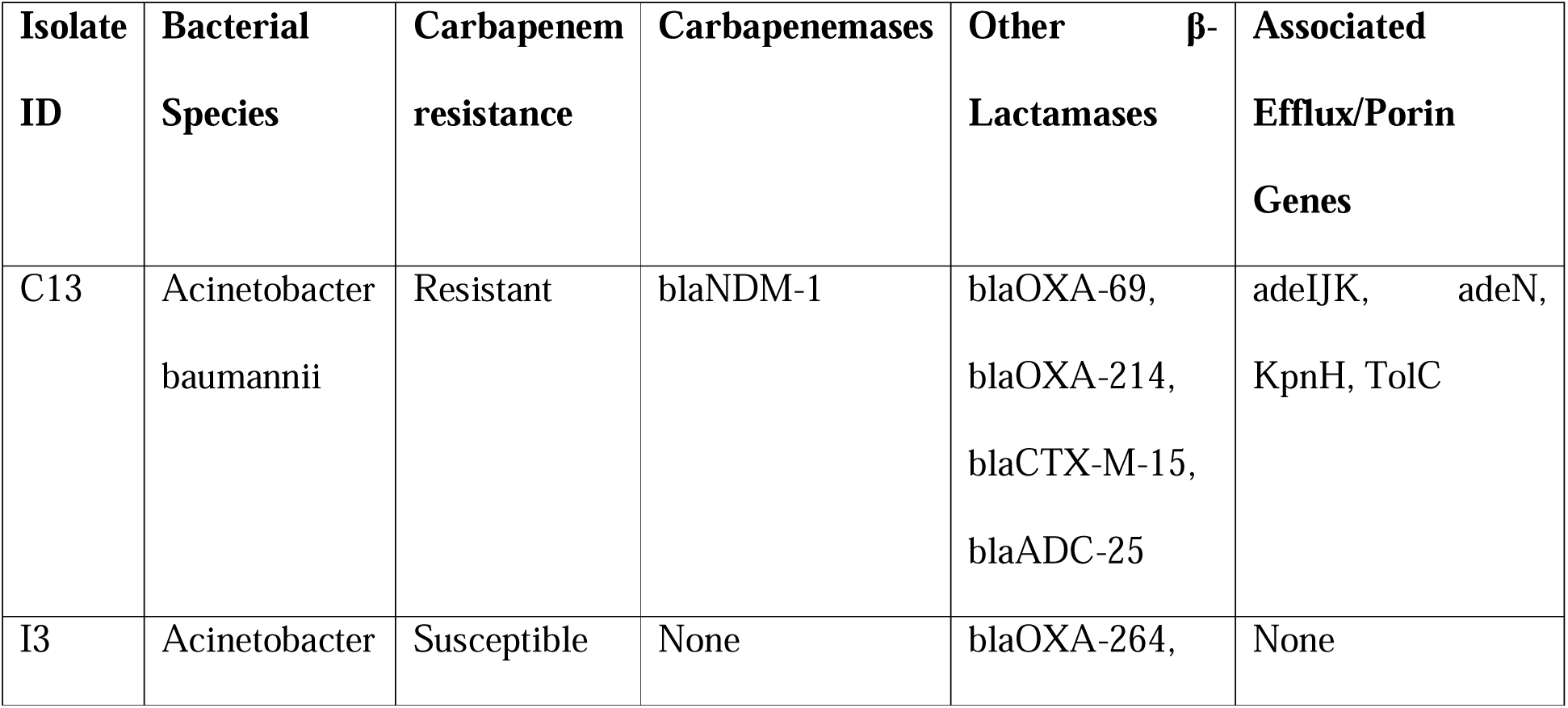
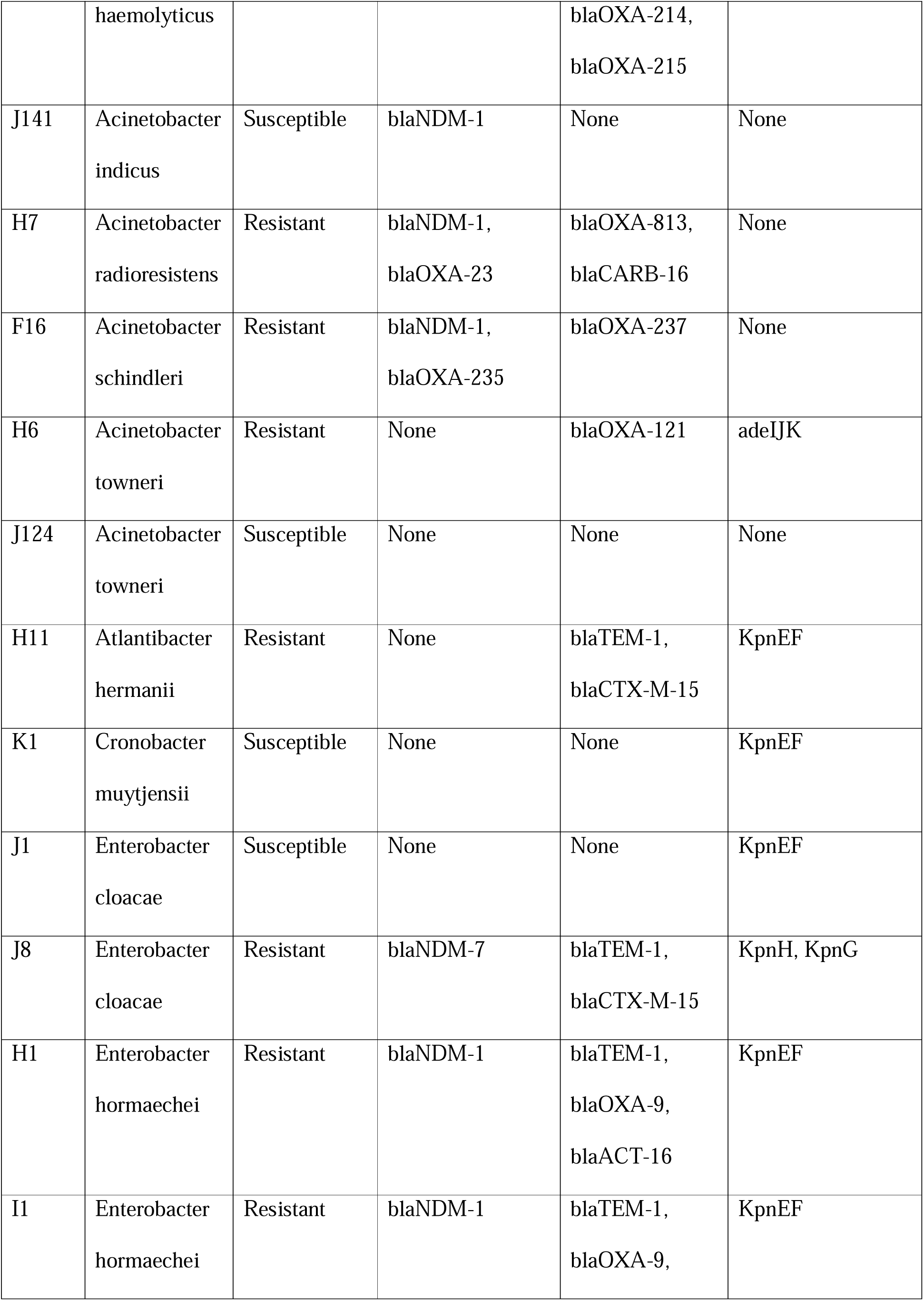
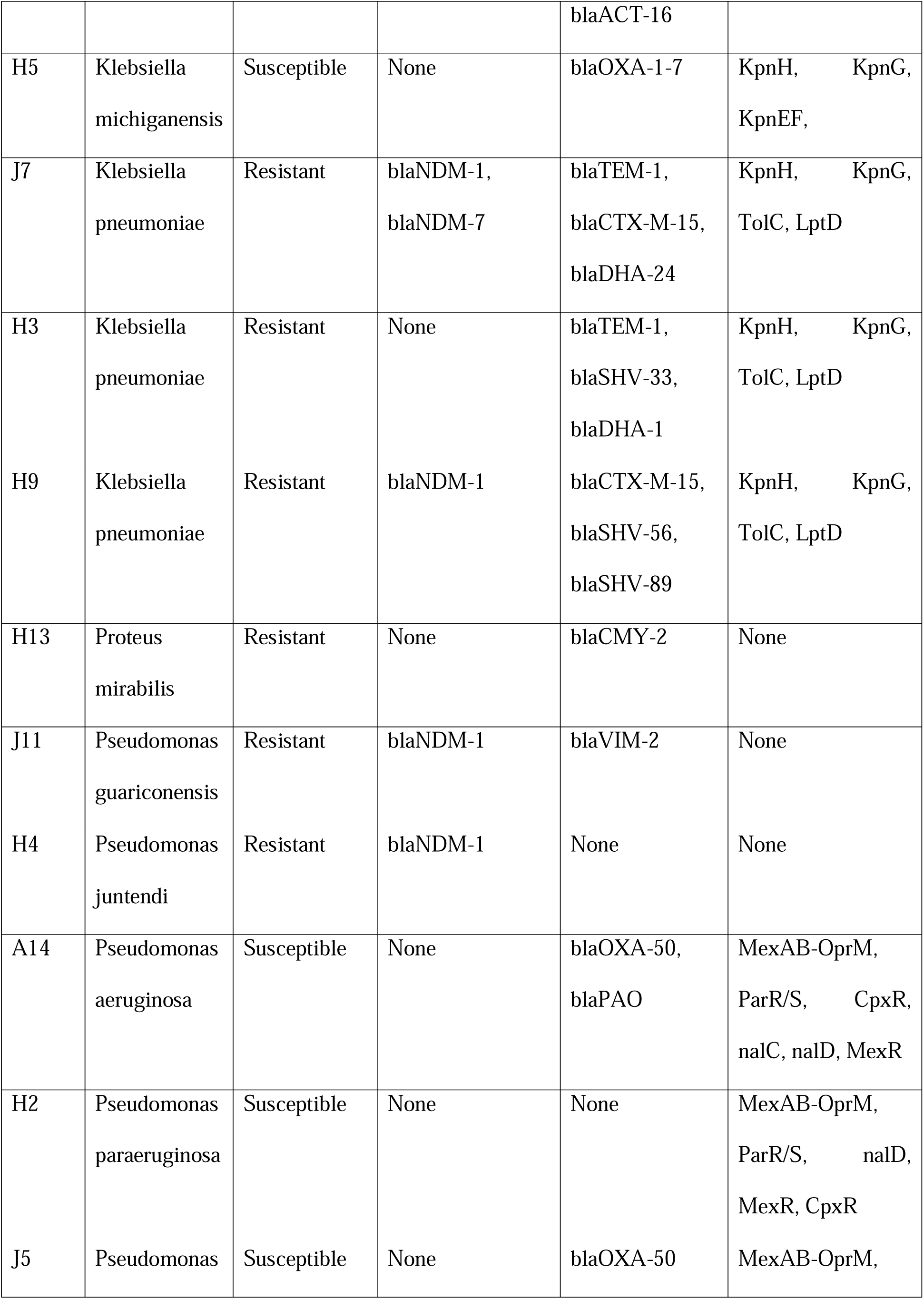
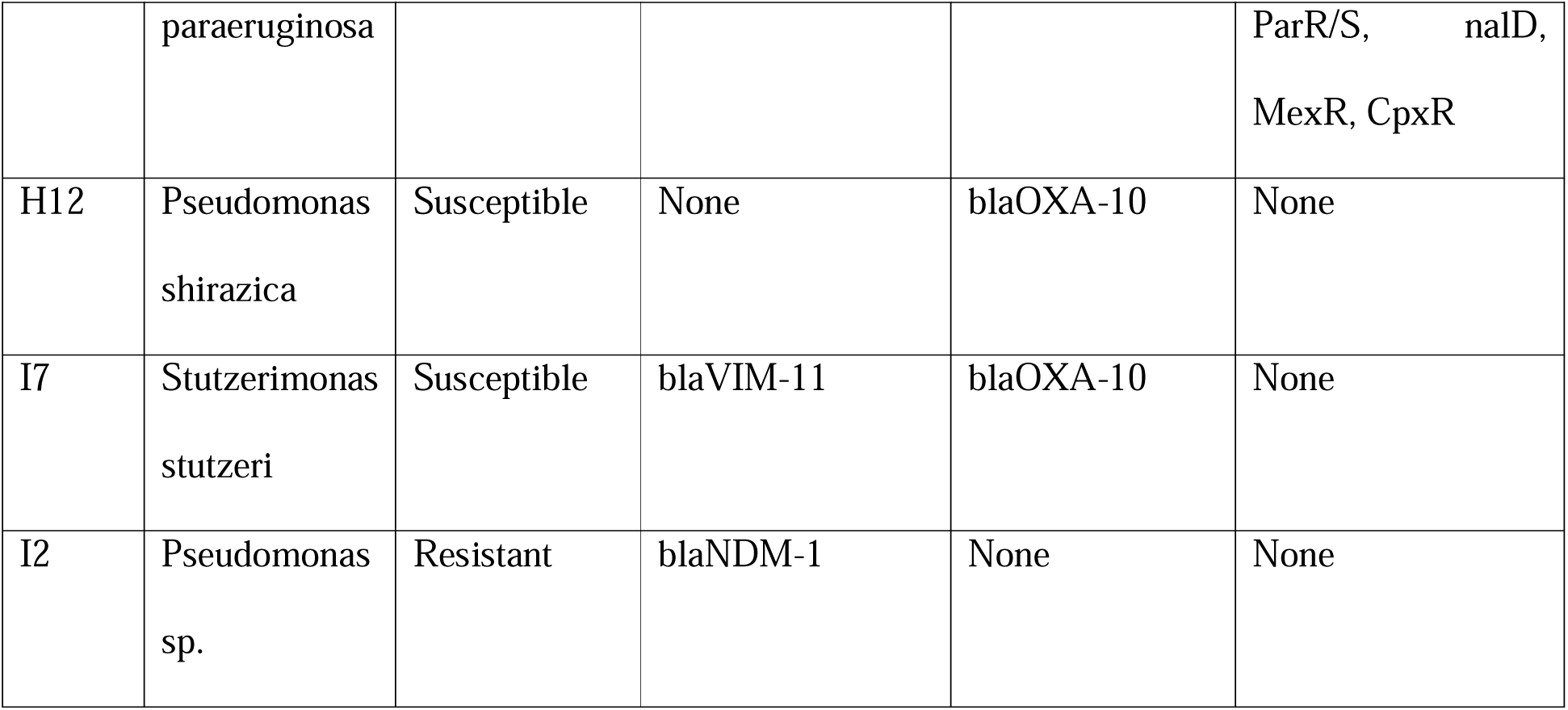
Genomic and phenotypic correlation of Carbapenems resistance.

In addition to carbapenemase genes, several other resistance determinants were detected, including efflux pump and regulatory genes such as adeIJK, adeN, CpxR, gyrA (T83I), KpnG, KpnH, LptD, MexABPQRYZ, nalC (A186T, G71E), nalD, opmE, OprM, ParR, ParS, and TolC.

Isolates of *Acinetobacter haemolyticus*, *Pseudomonas shirazica*, *Klebsiella michiganensis*, *Cronobacter muytjensii*, along with single isolates of Enterobacter cloacae and Acinetobacter towneri, notably did not display phenotypic carbapenem resistance nor did they carry any known carbapenem resistance genes.

Interestingly, one isolate of Atlantibacter hermanii exhibited phenotypic resistance to all three carbapenems (ertapenem, imipenem, and meropenem), yet no known carbapenemase genes were detected. In contrast, a Stutzerimonas stutzeri isolate carried the blaVIM-11 gene but remained phenotypically susceptible to carbapenems.

## Discussion

The presence of antibiotic resistant bacteria in non-clinical environments, such as in the air, wastewater, agriculture and water bodies acts as environmental reservoirs that facilitate the persistence and dissemination AMR. These environments act as “melting pots” where resistance genes can emerge, be maintained, and spread across microbial communities. Opportunistic pathogens such as Acinetobacter spp. and Pseudomonas spp. exhibit remarkable adaptability, frequently acquiring multidrug resistance under selective pressures. These resistant strains may be transported into external environments or may arise de novo outside hospitals, driven by the co-occurrence of antibiotic residues, biocides, and heavy metals. Such contaminants exert selective pressure that favors resistant populations even in the absence of direct antibiotic exposure, thereby sustaining and amplifying the environmental dimension of AMR ^26^.

Carbapenems are important group of broad-spectrum antibiotics that are listed as reserve group of antibiotics by WHO, meant to be considered as last resort option for multidrug resistant infections ^27^. There is an increase in carbapenem-resistant organisms especially GNB worldwide, and particularly in India where high burden of CRGs are reported. In a two-year study conducted by Lathakumari RH et al in tertiary care centre in South India, a high burden of carbapenem resistance in crucial Gram-negative isolates namely *Acinetobacter baumannii* and *Klebsiella pneumoniae* was observed ^28^. A prospective study in North India on AMR in clinical specimens showed high resistance rate in carbapenems-resistant A. baumanii ^29^. Systematic review and meta-analysis of studies on AMR from Eastern and North-Eastern states of India also revealed a high resistance to carbapenems in GNB ^30^. Such reports in literature prove beyond doubt the presence of high burden of CRGs in Indian hospital and clinical samples. However, from one health perspective, understanding the burden of CRGs in the environment outside hospitals and health care facilities is equally important. Untreated or inadequately treated hospital and urban sewage, together with animals and ready to eat vegetables, have been identified as important sources of carbapenem resistant organisms in the environment ^31^. However, the possibility of disposed solid waste, especially biomedical waste from hospitals and diagnostic and research laboratories as a potential source of AMR transmission to the outside environment has not been addressed due to lack of such studies in the literature. With an aim to fulfil this gap, the present study aimed to identify the phenotypic and genotypic characteristics of airborne CRGs isolated from BMW site in Gujarat, India.

In this study, 26 CRBs isolated from the BMW site belonged to 3 families, namely the Moraxellaceae (07 isolates), Enterobacteriaceae (11 isolates) and Pseudomonadacea (08 isolates). Although these bacteria were environmental isolates, they were of immense clinical relevance as they showed MDR phenotype, including resistance to multiple carbapenems. The importance of these families as potential reservoir of ARGs is also highlighted by a Chinese study done in the municipal landfill sites. It reported that 72.2 % of ARG bacteria isolated from 22 municipal landfills sites belonged to these three families, in addition to Xanthomonadaceae ^32^. Non-clinical species function as environmental reservoirs of antimicrobial resistance, where contamination and biological stress drive the emergence of new resistant strains ^33^. In our study, more than half of the isolates from all the 3 families showed carbapenem resistance and harboured either carbapenemase or/and other beta lactamases as well as associated efflux/porin genes.

The phenotypic profile of the 26 isolates revealed 58% isolates to be resistant to carbapenems while the remaining 42% showed a susceptible profile. The genotypic profiling of the 26 isolates offers interesting insights. blaNDM was found to be the resistant gene present in 12 isolates across all the three families with the variant blaNDM-1 being predominant one (11 of 12 isolates), while blaNDM-7 was present in two isolates, either alone or co-existed with blaNDM-1. Bacteria carrying blaNDM-1 are widely distributed with four main species, such as Klebsiella sp., Escherichia sp., Acinetobacter sp. and Pseudomonas sp. ^34^. A previous study on the genetic analysis of A. baumanii isolates from clinical samples collected in 2005 in South Indian hospital demonstrated that the blaNDM-1 gene emerged in Acinetobacter species before it spread to Enterobacteriaceae ^35^. In our study, different species of Acinetobacter including A. baumanii, and A. indicus harboured blaNDM-1 gene, while in A. radioresistens and A. schindeleri, blaOXA-23 and blaOXA-235, respectively co-existed with blaNDM-1. In addition to Acinetobacter sp., E. cloacae, E. hormaechei, K. pneumoniae, P. gauriconensis, and P. juntendii, also showed the presence of the blaNDM gene.

Acinetobacter baumanii is a common nosocomial pathogen, often isolated from the clinical samples and hospital environment and carries blaNDM-1 gene ^36^. A. baumanii is also reported in the waste water and natural water environments ^37,38^. Similarly, E cloaece and E hormaechi, members of Enterobacteriaceae cloacae complex and K pneumonia are frequently present in the clinical settings ^39–42^. Isolation of these species from the BMW site is not unexpected, but alarming given that they are opportunistic pathogens and apart from carbapenem-resistant gene, harbours other beta lactamases and associated efflux/porin genes. Other isolates, namely Acinetobacter radioresistens and Acinetobacter schindeleri, though not as abundant as the above strains, are reported to cause opportunistic infections under co-morbid conditions ^42,43^. Acinetobacter radioresistens can resist 4-8x higher γ-radiation than other *Acinetobacter* spp. and can spread OXA-23 carbapenemase, thereby conferring carbapenem resistance ^44^. We also observed the presence of blaNDM-1 gene in P gauriconesis, P juntendi and S stutzeri isolates, which are less frequent and recently reported strains from clinical samples ^45,46^. This existence of resistant genes may be the result of horizontal transfer.

A towneri, Atlantibacter hermanii, K pneumonia and P mirabilis showed phenotypic carbapenem resistance in the absence of carbapenem-resistant genes. However, they showed the presence of other beta lactamases and/or efflux/porin genes, which may have conferred carbapenem resistance^47^. Despite the presence of carbapenem-resistant genes, blaNDM-1 and blaVIM-11 in A. indicus and S. stutzeri, respectively, the isolates remained susceptible to carbapenems. It is interesting to note that A. indicus lacked other beta lactamases or efflux/porin genes, while blaOXA-10 was present in S stutzeri. Also, A. indicus-like strains, carrying blaOXA-23 on transposable elements and displaying carbapenem resistance have been isolated from the cattle ^48^. S. stutzeri, also known as *P. stutzer*, isolated from porcine faeces showed a novel-type plasmid, co-harbouring blaVIM-2 and tmexCD1-toprJ ^32,49^.

The findings of the present study support that environmental monitoring of AMR at BMW sites is crucial in AMR surveillance, as inappropriate use of antibiotics and improper management of healthcare waste can have direct and indirect health consequences for healthcare personnel, waste site workers, the community, and the environment. The spread of airborne antibiotic-resistant bacteria at BMW disposal sites can occur due to the aerosolization of antibiotic-resistant bacteria (ARB) and ARGs from the infectious hospital waste, often contributed by improper packaging of the biohazardous waste (red) bags, bioaerosols generation during from transportation from hospitals to the BMW disposal sites by the collection vehicles, and deposition of bioaerosols at the indoor and outdoor environments of the BMW sites ^50^. The biophysical and anthropogenic factors ^51^, including climatic and meteorological conditions ^52^, disturbances induced by transport and wind, animals, water action, cleaning and sanitation measures at hospitals and other collection sites, inadequate segregation, packaging, disposal of wastes from hotspots like healthcare facilities ^53^, and natural human activities at the BMW disposal sites, such as breathing, talking, coughing, sneezing, and shedding of skin cells can spread bioaerosols carrying ARBs and ARGs. The diversity of ARBs in bioaerosols at the BMW sites depends on the level of human activities and the efficiency of ventilation systems at these sites ^54^.

## Data availability

The whole genome sequencing data generated and analysed in this study have been deposited in the NCBI BioProject database and are publicly accessible under accession number PRJNA1085497. (URL: https://www.ncbi.nlm.nih.gov/bioproject/1085497)

## Acknowledgements

The authors thank Mrs. Jyotsana Verma and Mr. Ajay Kumar for help in.

## Transparency declarations

The authors declare that they have no conflict of interest related to this study.

